# Spinal muscular atrophy diagnosis and carrier screening from genome sequencing data

**DOI:** 10.1101/19006635

**Authors:** Xiao Chen, Alba Sanchis-Juan, Courtney E French, Andrew J Connell, Isabelle Delon, Zoya Kingsbury, Aditi Chawla, Aaron L Halpern, Ryan J Taft, NIHR BioResource, David R Bentley, Matthew ER Butchbach, F Lucy Raymond, Michael A Eberle

## Abstract

**Purpose:** Spinal muscular atrophy (SMA), caused by loss of the *SMN1* gene, is a leading cause of early childhood death. Due to the near identical sequences of *SMN1* and *SMN2*, analysis of this region is challenging. Population-wide SMA screening to quantify the *SMN1* copy number (CN) is recommended by the American College of Medical Genetics.

**Methods:** We developed a method that accurately identifies the CN of *SMN1* and *SMN2* using genome sequencing (GS) data by analyzing read depth and eight informative reference genome differences between *SMN1/2*.

**Results:** We characterized *SMN1/2* in 12,747 genomes, identified 1568 samples with *SMN1* gains or losses and 6615 samples with *SMN2* gains or losses and calculated a pan-ethnic carrier frequency of 2%, consistent with previous studies. Additionally, 99.8% of our *SMN1* and 99.7% of *SMN2* CN calls agreed with orthogonal methods, with a recall of 100% for SMA and 97.8% for carriers, and a precision of 100% for both SMA and carriers.

**Conclusion:** This *SMN* copy number caller can be used to identify both carrier and affected status of SMA, enabling SMA testing to be offered as a comprehensive test in neonatal care and an accurate carrier screening tool in GS sequencing projects.

## Introduction

Spinal muscular atrophy (SMA), an autosomal recessive neuromuscular disorder characterized by loss of alpha motor neurons, causes severe muscle weakness and atrophy presenting at or shortly after birth^1,2^. SMA is the leading genetic cause of infant death after cystic fibrosis^3^. The incidence of SMA is 1 in 6000-10,000 live births, and the carrier frequency is 1:40-80 among different ethnic groups^4–7^. Four clinical types of SMA are classified based on age of onset and severity of the disease^1^: very weak infants unable to sit unsupported (Type I), weak sitters but unable to stand (Type II), ambulant patients with weaker legs than arms (Type III) and adult onset SMA (Type IV). Early detection of SMA can be critical for long term quality of life due to the availability of two early treatments, Nusinersen^8^ and Zolgensma^9^, which have received FDA approval for the amelioration of SMA symptoms.

The disease causing gene, *SMN1*, together with its paralog *SMN*2, reside in a ∼2Mb region on 5q with a large number of complicated segmental and inverted segmental duplications. *SMN2* is 875kb away from *SMN1* and is created by an ancestral gene duplication that is unique to the human lineage^10,11^. The genomic region around *SMN1/2* is subject to unequal crossing-over and gene conversion, resulting in variable copy numbers (CNs) of *SMN1* and *SMN2*^7,12^. Importantly, *SMN2* has >99.9% sequence identity to *SMN1* and one of the base differences, NM_000344.3:c.840C>T in exon 7, has a critical functional consequence. By interrupting a splicing enhancer, c.840T promotes skipping of exon 7, resulting in the vast majority of *SMN2*-derived transcripts (70-85%, depending on tissue^13^) being unstable and not fully functional. Approximately 95% of SMA cases result from biallelic absence of the functional c.840C nucleotide^14^ caused by either a deletion of *SMN1* or gene conversion to *SMN2* (c.840T). In the remaining 5% of SMA cases, patients also have other pathogenic variants in *SMN1*^15^. *SMN2* can produce a small amount of functional protein, and the number of *SMN2* copies in an individual modifies the disease severity and is highly correlated with the clinical types described above^16^.

Due to the high incidence rate and disease severity, population-wide SMA screening is recommended by the American College of Medical Genetics^17^. The utility of population-wide carrier screening has been demonstrated in pilot studies^18^. The key to screening for SMA is: 1) determining the copy number of *SMN1* for SMA diagnosis and carrier testing and 2) determining the copy number of *SMN2* for clinical classification and prognosis. Traditionally, SMA testing and carrier testing are done with polymerase chain reaction (PCR) based assays, such as quantitative PCR (qPCR)^19^, multiplex ligation-dependent probe amplification (MLPA)^20,21^ and digital PCR^12,22^. These methods primarily determine the copy number of *SMN1* based on the c.840C>T site that differs between *SMN1* and *SMN2*.

With recent advances in next-generation sequencing (NGS), it is now possible to profile a large number of genes or even the entire genome at high throughput and in a clinically relevant timeframe. Driven by these advances, many countries are undertaking large scale population sequencing efforts^23–25^ wherein testing for rare genetic disorders including carrier status will be one of the primary drivers. Demonstrating that GS can meet or exceed the performance of PCR-based SMA tests would indicate that both current and future precision medicine initiatives could leverage genome data for population-level screening. Replicating the current SMA testing regime poses a problem for high throughput GS due to the almost perfect sequence identity between *SMN1* and *SMN2*. Furthermore, it is thought that frequent gene conversion between *SMN1* and *SMN2* leads to hybrid genes. These challenges demand an informatics method specifically designed to overcome the difficulties of this region.

To date, two NGS-based tests for SMA carrier detection have been reported^26,27^. Larson et al.^26^ used a Bayesian hierarchical model to calculate the probability that the fraction of *SMN1*-derived reads is equal to or smaller than 1/3 at three base differences between *SMN1* and *SMN2*. Since this method does not perform copy number calling, it is not an ideal solution for carrier screening. Conversely, Feng et al.^27^ developed a copy number caller for both *SMN1* and *SMN2* based on targeted sequencing data that closely mimics the current PCR-based method. Their method is designed for targeted sequencing and thus requires specialized normalization that limits their method to one assay at one site. More importantly, the relatively low depths of GS (∼30x) cannot provide reliable bp-resolution CN calls (see Figure S1) and since this method relies on a single locus, it is not amenable to accurate CN calls with GS. Precision-medicine initiatives will need a way to diagnose and detect carriers of SMA from GS data.

Here, we report a novel method that detects the CN of both *SMN1* and *SMN2* using GS data. While most conventional assays only test for the absence of c.840C as a proxy for the “exon 7 deletion” (either a true deletion or as a product of gene conversion with *SMN2*), here we describe a tool that can more fully characterize the variability in the region including: 1) DNA deletions, including both whole gene deletion/duplication and a partial deletion of a region that includes exons 7 and 8; and 2) small variant detection including the NM_000344.3:c.*3+80T>G (also referred to as g.27134T>G in literature) SNP that is correlated with “silent” carriers of SMA (two copies of *SMN1* on the same haplotype)^28^. To demonstrate the accuracy of this method, we compared CN calls using digital PCR and MLPA with our GS-based calls and showed a concordance of 99.8% for *SMN1* and 99.7% for *SMN2*. Additionally, we applied this method to 2504 unrelated samples from the 1000 Genomes Project^29^ (1kGP) and 10,243 unrelated samples from the NIHR BioResource Project^30^ to report on the population distributions of *SMN1* and *SMN2* copy numbers. The carrier frequencies for SMA using this method agreed with those reported by previous PCR-based studies^5,6^. In addition to demonstrating the accuracy of our method to quantify variants in the *SMN* region, we also highlight the importance of using ethnically diverse populations when developing novel informatic methods to resolve difficult clinically relevant regions of the genome.

## Materials and methods

### Samples and data processing

Samples validated using digital PCR were procured from the Motor Neuron Diseases Research Laboratory (Nemours Alfred I. duPont Hospital for Children) collection and were generated from cell lines as described previously^12,31^. Historical patient samples with known SMA or carrier status measured by MLPA were obtained from Cambridge University. GS was performed on 73 samples with digital PCR results, 45 samples with MLPA results, and 12,747 population samples from the 1000 Genomes Project^29^ (1109 of which have MLPA calls), the NIHR BioResource Rare Diseases project^30^ and the Next Generation Children (NGC) project^32^. The sequencing and processing of this data was done using a variety of sample preparation methods, Illumina sequencers, and read aligners. A full description of this data can be found in the Supplementary Methods.

### *SMN* copy number analysis by orthogonal methods

*SMN1* and *SMN2* CNs were measured for 73 samples using the QuantStudio 3D Digital PCR System (Life Technologies) using allele-specific exon 7 probes as described previously^12^. *SMN1* and *SMN2* copy numbers were normalized against those for *RPPH1* (*RNase P*). MLPA CN calls for 1109 of the 1kGP samples were available from Vijzelaar et al.^33^. 45 historical patient samples were previously tested in a clinically accredited laboratory by MLPA for SMN1/2 exon7 and exon8 copy number (SALSA MLPA P060 SMA Carrier probemix, MRC-Holland). Additionally, 2 samples from the Next Generation Children project were confirmed using MLPA.

### Copy number calling for intact and truncated *SMN*

Two common CNVs involve the *SMN1* and *SMN2* loci, the whole-gene CNV and a partial gene deletion of exons 7 and 8 (*SMN2*Δ*7-8*)^33,34^ (see Results). We first count reads that align to either *SMN1* or *SMN2*. Read counts in a 22.2kb region encompassing Exon 1 to Exon 6 are used to calculate the total *SMN* (*SMN1, SMN2* and *SMN2*Δ*7-8*) CN, and read counts in the 6.3kb region including Exon 7 and Exon 8 are used to calculate the CN of intact *SMN* (*SMN1* and *SMN2*). Read counts are normalized and converted into copy numbers using a one-dimensional mixture of Gaussian distributions (see Supplementary Information). The copy number of truncated *SMN* (*SMN2*Δ*7-8*) is derived by subtracting the intact *SMN* CN from total *SMN* CN.

### Genotyping *SMN1/2* copy number using differentiating bases

We call the number of chromosomes carrying the *SMN1* and *SMN2* bases by combining the total *SMN* CN with the read counts supporting each of the gene-specific bases. At each *SMN1/2* differentiating base (Table S1), based on the called copy number of intact *SMN*, the caller iterates through all possible combinations of *SMN1* and *SMN2* copy numbers and derives the combination that produces the highest posterior probability for the observed number of *SMN1* and *SMN2* supporting reads. The *SMN1* CNs called at single bases are then combined to make the aggregate *SMN1* CN call based on a consensus rule (see Supplementary Methods). In addition to calling the CN of bases that are specific to either *SMN1* or *SMN2*, this method can be applied to variant positions to identify the copy number of bases known to be specific to one of the two genes, e.g. c.*3+80T>G as described in Results.

## Results

### Common CNVs involving the *SMN1*/*2* loci

While existing PCR- or NGS-based methods focus primarily on the c.840C>T site, we adopted a copy number approach based on the sequencing data from the full genes. We examined the read depth across the ∼30kb homologous region harboring *SMN1* and *SMN2* genes in 1kGP samples (Figure 1A). The depth profile shows that this entire region can be deleted or duplicated in the population. The exact breakpoints of this CNV are expected to vary from sample to sample due to the extensive sequence homology within and beyond this region and can only be resolved in high resolution with long read sequencing. We restricted our CN analysis to the (∼30kb) regions that include the *SMN* genes (*SMN1* or *SMN2*).

**Figure 1.**
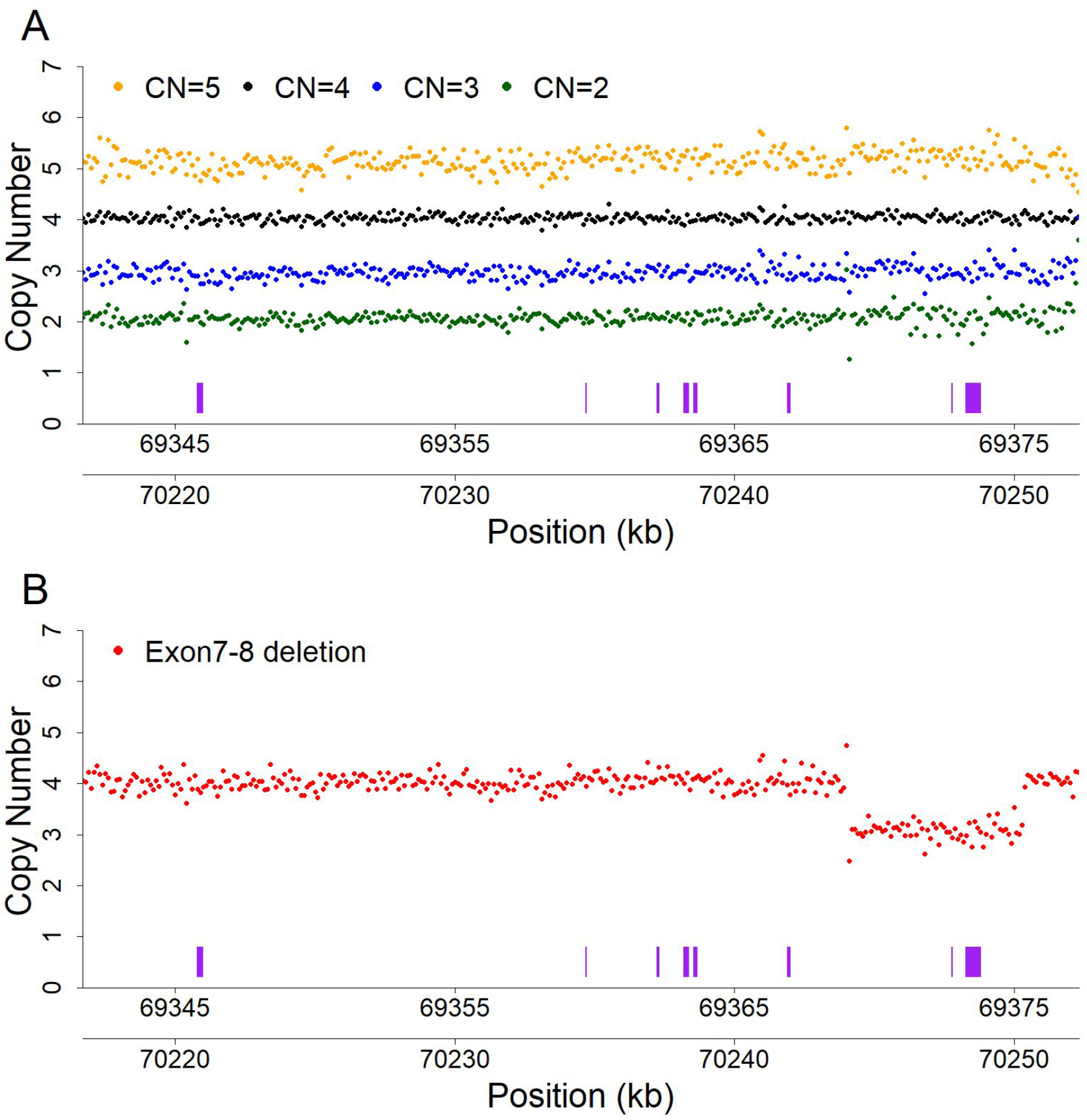
Common CNVs affecting the *SMN1*/*2* loci. **A**. Depth profiles across the *SMN1/SMN2* regions. Samples with a total *SMN1*+*SMN2* copy number of 2, 3, 4 and 5 (derived from average read depth in the two genes) are shown as green, blue, black, and orange dots, respectively. Depth from 50 samples are summed up for each CN category. Each dot represents normalized depth values in a 100bp window. Read counts are calculated in each 100bp window, summing up reads from both *SMN1* and *SMN2*, and normalized to the depth of wild-type samples (CN=4). The *SMN* exons are represented as purple boxes. The two x axes show coordinates (hg19) in *SMN1* (bottom) and *SMN2* (upper). **B**. Depth profiles aggregated from 50 samples carrying a deletion of exons 7 and 8 are shown as red dots. Read depths are calculated in the same way as in (A).

In addition to whole gene CNVs, we also found a 6.3kb partial gene deletion encompassing both exons 7 and 8 (Figure 1B, Figure S2) that was recently described in another study^34^. The sequences at the breakpoint are identical between *SMN1* and *SMN2*, so this deletion occurs at either chr5: 70244114 - 70250420 in *SMN1* or chr5: 69368689 - 69375000 in *SMN2* (Figure S2, hg19). However, about 500bp downstream from the breakpoint that defines the end of this deletion there are three base differences between the *SMN1* and *SMN2* loci (70250881A>69375425C, 70250981A>69375525G, 70250991A>69375535G). Among the 1kGP samples that contain this deletion, we identified 245 read pairs from 237 samples where one spanned the breakpoint and the mate spanned at least two of the three *SMN*-differentiating bases. Analysis of these read pairs revealed that 100% were consistent with the deletion occurring on the *SMN2* sequence background. We named this truncated form of *SMN2* “*SMN2*Δ*7-8*”. Since both exons 7 and 8 are deleted, *SMN2*Δ*7-8* most likely has limited or no biological function. Therefore, *SMN2*Δ*7-8* is an important variant that any *SMN* CN caller should take into account.

After searching for anomalous read pairs in the 1kGP samples, we did not identify any other common CNVs in the *SMN* region. Combining this information together, we called CNs of the *SMN* genes to specifically identify the number of intact and truncated forms by dividing the genes into two regions: the 6.3kb region that includes exons 7-8 and the 22.2kb region that includes exons 1-6. The CN calculated from the exons 7-8 region provided the number of intact *SMN* genes. Samples with *SMN2*Δ*7-8* have a higher CN call from the exon 1-6 region compared to the CN call from the exon 7-8 region, and this difference represents the CN of *SMN2*Δ*7-8*. Figure 2 shows the results of this calculation for 12,747 samples where we identified 2144 instances of *SMN2*Δ*7-8* including 140 samples with two copies of *SMN2*Δ*7-8* and one sample with three copies of *SMN2*Δ*7-8*.

**Figure 2.**
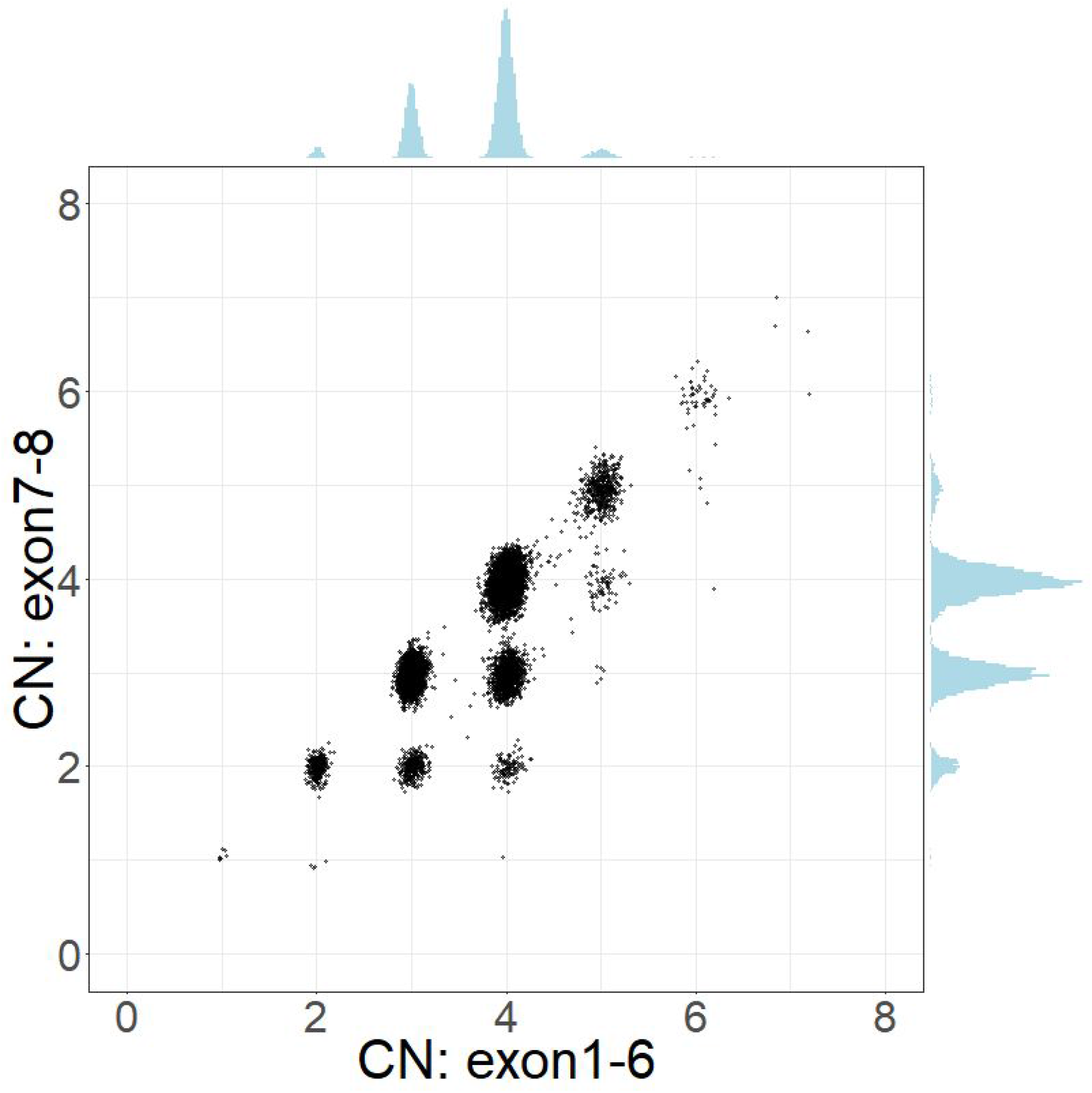
Scatterplot of total *SMN* (*SMN1, SMN2* and *SMN2*Δ*7-8*) copy number (x axis, called by read depth in Exon 1-6) and intact *SMN* copy number (y axis, called by read depth in Exon 7-8).

### *SMN1*/2 CN calls using differentiating bases

We called the CNs of *SMN1* and *SMN2* at the 16 base difference sites between *SMN1* and *SMN2* (see Methods, Table S1) in 1kGP samples, and compared the CN calls for each position with the CN calls at the splice variant site (Figure 3A, Figure S3). There was a notable difference between the concordance of calls in the African and non-African populations (Figure 3A). Excluding the African samples, there were 13 sites that had high (>85%) CN concordance with the splice variant site. Conversely, for the African samples there were only seven sites that had high CN concordance with the splice variant site, and the concordance values were lower than in non-African populations. This is consistent with within-gene variation at many of these positions in the African population. We selected the splice variant site and the seven positions that were highly concordant with the splice variant site in both African and non-African populations to make *SMN1* and *SMN2* CN calls based on the consensus of CN calls at the selected sites (see Supplementary Information for the detailed rules and in-depth analysis on the variability of the 16 sites).

**Figure 3.**
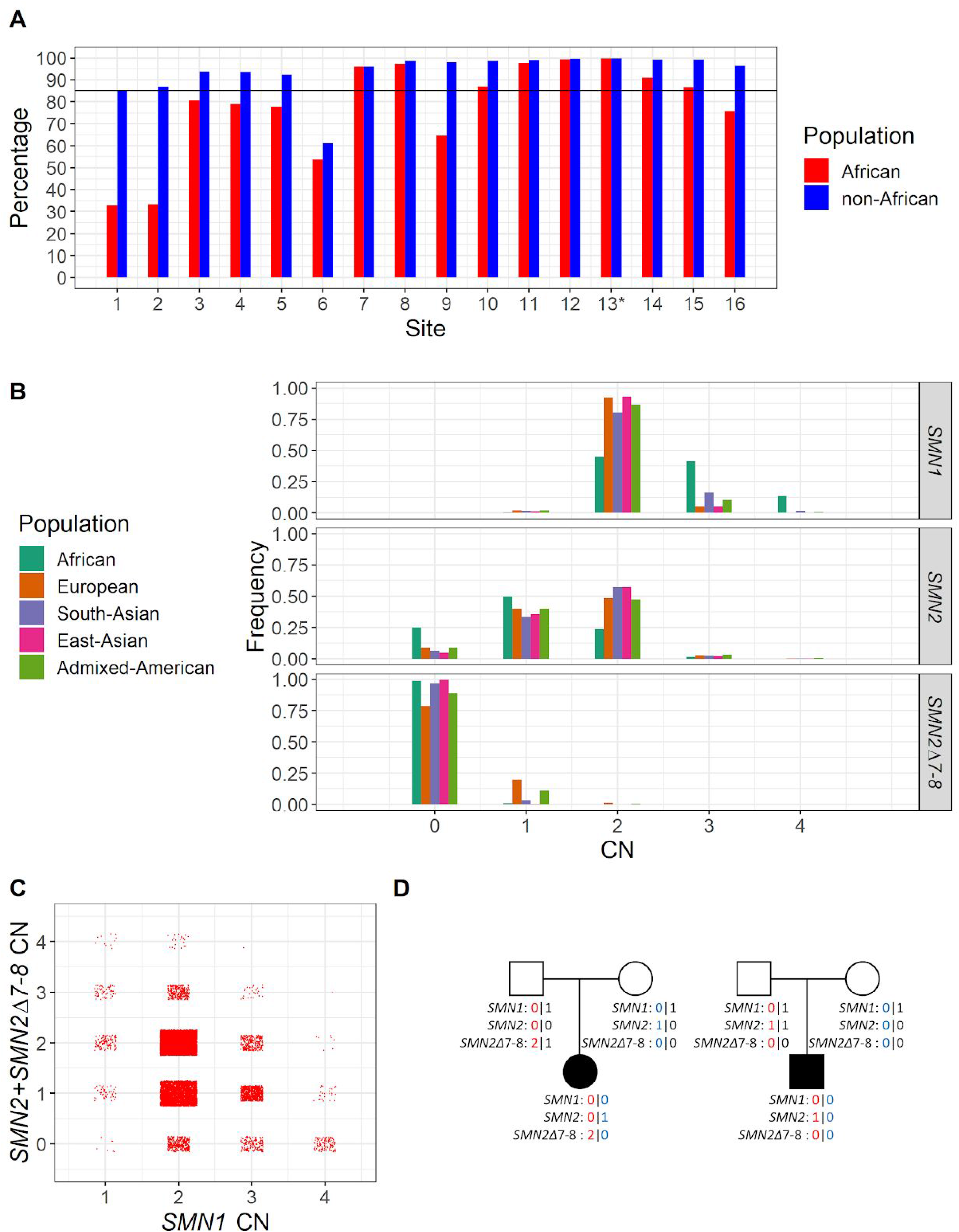
Distribution of *SMN1*/*SMN2*/*SMN2*Δ*7-8* copy numbers in the population. **A**. Percentage of samples showing CN call agreement with c.840C>T across 16 *SMN1-SMN2* base difference sites in African and non-African populations. Coordinates of these 16 sites are given in Table S1 and site 13* is the c.840C>T splice variant site. The black horizontal line denotes 85% concordance. **B**. Histogram of the distribution of *SMN1, SMN2* and *SMN2*Δ*7-8* copy numbers across five populations in 1kGP and the NIHR BioResource cohort. **C**. *SMN1* CN vs. total *SMN2* CN (intact *SMN2* + *SMN2*Δ*7-8*). **D**. Two trios with an SMA proband detected by the caller and orthogonally confirmed in the Next Generation Children project cohort. CNs of *SMN1, SMN2* and *SMN2*Δ*7-8* are phased and labeled for each member of the trios.

### Validation of the *SMN* copy number caller

After developing this method, we tested its accuracy against results from orthogonal methods. We sequenced 73 samples with known *SMN1* and *SMN2* CNs measured by digital PCR; 45 samples with known results measured by MLPA; and also compared our CN calls with MLPA calls published by Vijzelaar et al.^33^ on 1109 samples from the 1kGP. This validation data include 64 SMA probands, 45 SMA carriers and 1118 samples with an *SMN1* CN larger than 1. Our *SMN1* CN calls agreed with digital PCR results in 99.8% of the samples, the *SMN2* CN calls agreed in 99.7% of the samples, and *SMN2*Δ*7-8* calls agreed in 100% of the samples (Table 1, Table S2). Combined, our recall is 100% for SMA (*SMN1* CN=0, 64/64), 97.8% for carriers (*SMN1* CN=1, 44/45, the one missed carrier does not show read evidence supporting the carrier status, see below and Supplemental Information), and our precision is 100% for both SMA (1160/1160) and carriers (1179/1179). We also analyzed the consistency of *SMN1/SMN2*/*SMN2*Δ*7-8* CN calls in 258 trios from the Next Generation Children project cohort (see Methods). There is no Mendelian error in any of the calls (Table S3).

**Table 1.** Validation against samples with known *SMN1*/*2* CNs.

|  | CN by orthogonal method | Total | Concordant | Discordant | Agreement |
| --- | --- | --- | --- | --- | --- |
| *SMN1* | 0 | 64 | 64 | 0 | 100.0% |
|  | 1 | 45 | 44 | 1 | 97.8% |
|  | 2 | 897 | 897 | 0 | 100.0% |
|  | 3 | 174 | 174 | 0 | 100.0% |
|  | 4 | 43 | 43 | 0 | 100.0% |
|  | 6 | 1 | 0 | 1 | 0.0% |
|  | Total | 1224 | 1222 | 2 | 99.8% |
| *SMN2* | 0 | 117 | 117 | 0 | 100.0% |
|  | 1 | 466 | 465 | 1 | 99.8% |
|  | 2 | 541 | 539 | 2 | 99.6% |
|  | 3 | 60 | 60 | 0 | 100.0% |
|  | 4 | 9 | 8 | 1 | 88.9% |
|  | Total | 1193 | 1189 | 4 | 99.7% |
| *SMN2Δ7-8* | 0 | 1089 | 1089 | 0 | 100.0% |
|  | 1 | 80 | 80 | 0 | 100.0% |
|  | 2 | 4 | 4 | 0 | 100.0% |
|  | Total | 1173 | 1173 | 0 | 100.0% |

Further inspection of the five discrepant samples does not show evidence of errors in our calls except in one sample (see Supplemental information). The validation for the 73 and 45 samples was done using two sets of probes targeting Exon 7 and Exon 8 of *SMN1* and *SMN2*, while the validation for the 1109 samples used a newer version of the MLPA probemix which includes an additional 17 probes used to determine the *SMN1*+*SMN2* CN in Exon1-6 and Exon7-8, which helped refine the individual *SMN1* and *SMN2* CN calls. Correspondingly our concordance is significantly higher (p-value = 0.009223 for *SMN1*, 2.651e-05 for *SMN2*, Fisher’s exact test) measured against the newer version of MLPA (100% concordance for *SMN1* and *SMN2*) than other methods (98.3% for *SMN1* and 95.4% for *SMN2*).

### Copy number of *SMN1, SMN2* and *SMN2*Δ*7-8* by population

Given the high accuracy demonstrated by our validation against orthogonal methods, we next applied this method to high depth (all samples sequenced to at least 30x) GS data from 12,747 unrelated samples from the 1kGP and the NIHR BioResource Project (Table S4). We analyzed the CN distributions by population (Europeans, Africans, East Asians, South Asians and admixed Americans consisting of Colombians, Mexican-Americans, Peruvians and Puerto Ricans). Figure 3B shows the histogram of the number of individuals with different CNs of intact *SMN1*, intact *SMN2* and *SMN2*Δ*7-8*. The distributions are similar between the 1kGP samples and the NIHR BioResource samples (Figure S4, Table S5). In general, individuals have more copies of *SMN1* than *SMN2*. The most common combinations of *SMN1/SMN2* copy number are 2/2 (44.9%) and 2/1 (33.4%). The variability of *SMN1* copy number is much lower than that of *SMN2* copy number in most populations, while the Africans show higher variability in both *SMN1* and *SMN2* CN. Conversely, 54.7% of Africans have three or more copies of *SMN1*, which is more than double what is observed in any of the other four populations (Figure 3B, Table 2). There is an inverse relationship between the copy number of *SMN1* and *SMN2* (Figure 3C, correlation coefficient −0.344, p-value < 2.2e-16). This observation is consistent with a mechanism where gene conversion occurs between *SMN1* and *SMN2*^35,36^. The observed higher *SMN1* CN relative to *SMN2* CN could be a result of a bias towards *SMN2*-to-*SMN1* conversion or selection against a low *SMN1* CN. Africans have significantly lower *SMN2* CN than the other populations (Wilcoxon test, p-value < 2.2e-16).

**Table 2.** *SMN1* CN and c.*3+80T>G frequency by population.

| Ethnicity | Total | *SMN1* CN=1 |  | *SMN1* CN=2 |  | *SMN1* CN=3 |  | *SMN1* CN=4 |  |
| --- | --- | --- | --- | --- | --- | --- | --- | --- | --- |
|  |  | Count | c.*3+80T>G+ | Count | c.*3+80T>G+ | Count | c.*3+80T>G+ | Count | c.*3+80T>G+ |
| African | 902 | 4 | 0<br>(0.0%) | 404 | 134<br>(33.17%) | 373 | 315<br>(84.45%) | 121 | 112<br>(92.56%) |
| European | 9648 | 212 | 0<br>(0.0%) | 8899 | 4<br>(0.04%) | 524 | 22<br>(4.2%) | 13 | 2<br>(15.38%) |
| South-Asian | 1199 | 20 | 0<br>(0.0%) | 965 | 1<br>(0.1%) | 195 | 5<br>(2.56%) | 19 | 1<br>(5.26%) |
| East-Asian | 593 | 8 | 0<br>(0.0%) | 552 | 1<br>(0.18%) | 33 | 1<br>(3.03%) | 0 | 0<br>(NA) |
| Admixed-American | 341 | 7 | 0<br>(0.0%) | 296 | 7<br>(2.36%) | 36 | 9<br>(25.0%) | 2 | 1<br>(50.0%) |

The number of SMA carriers identified across populations is summarized in Table 2 and Table S6. In 12,683 individuals with confident *SMN1/SMN2* CN calls, Europeans have the highest carrier frequency at 2.2%, followed by admixed Americans (2.05%), South Asians (1.67%) and East Asians (1.35%). Africans have the lowest carrier frequency (0.44%) by *SMN1* CN, and this carrier frequency does not include the 2+0 SMA silent carriers that are more common in Africans (see “Detection of silent carriers” below). The CN frequency distributions observed in this study are consistent with previous studies of *SMN1/SMN2* CN distribution in the general population^5,6^. In addition, we also report the frequency of the exon 7-8 deletion (*SMN2*Δ*7-8*) across populations. 21.2% of Europeans and 11.5% of admixed Americans have at least one copy of *SMN2*Δ*7-8*, while the frequency is lower in South Asians (3.35%), Africans (1.1%) and East Asians (0.34%).

In the Next Generation Children project cohort (see Methods), we identified SMA in two neonatal probands from trio analysis, which were confirmed independently. *SMN1, SMN2* and *SMN2*Δ*7-8* CNs are phased for each trio member (Figure 3D).

### Detection of “silent” carriers

The c.*3+80T>G SNP has been reported to be associated with the 2+0 SMA silent carrier status where one chromosome carries two copies of *SMN1* (either by *SMN1* duplication or gene conversion of *SMN2* to *SMN1*) and the other chromosome has no copies of *SMN1*^28^. Our method can also detect the presence of this SNP and thus can be used to screen for potential silent carriers. In the population samples, this SNP is differentially associated with two-copy and one-copy *SMN1* haplotypes and can be used to identify silent carriers (Table 2). We estimate the sensitivity improvement (with the biggest improvement from 70.5% to 91.8% in Africans) and residual risks for calling SMA carriers using the combination of *SMN1* CN and c.*3+80T>G SNP calling (see Supplementary information, Table S7, Table S8). One of our validation samples is a silent carrier and we correctly called *SMN1* CN=2 and the presence of c.*3+80T>G SNP (Table S2).

## Discussion

Due to the high sequence homology between *SMN1* and *SMN2*, the *SMN* region is difficult to resolve with both short and long read sequencing and thus far this important region has been excluded from standard GS analysis. Here, we demonstrate an algorithm that can resolve the CNs of *SMN1* and *SMN2* independently using short-read GS data, filling in an important gap in SMA diagnosis and carrier screening for precision medicine initiatives. Accurate measurement of *SMN1* and *SMN2* CNs is essential not only for the diagnosis of SMA but is also a prognostic indicator and the basis of therapeutic options^37^. *SMN2* CN has been used as a criterion for many clinical trials for SMA, including Nusinersen^8^ and Zolgensma^9^.

As a demonstration of this algorithm, we made CN calls for *SMN1* and *SMN2* using sequencing data from 12,747 samples covering five distinct subpopulations. We identified a total of 251 samples with *SMN1* losses (less than two copies) and 1317 with *SMN1* gains (more than two copies); 6241 samples with *SMN2* losses and 1274 with *SMN2* gains; 2144 samples carrying one or more copies of the truncated form *SMN2*Δ*7-8*. We cannot quantify the role that deletions, duplications or gene conversion play to drive the CN changes in this region but we see evidence supporting all three mechanisms including: 1) 3853 samples with total (*SMN1*+*SMN2*) CN<4 (deletions), 2) 670 samples with total CN>4 (duplications) and 3) a strong inverse correlation between the *SMN1* and *SMN2* CN (gene conversion, Figure 3C). Additionally, we identified a carrier frequency between 1:45 and 1:225 depending on ancestral population (Table 2). Comparing the CN frequencies by population shows that they are highly different and our per-population results agree with previous population studies^5,6^. While this provides qualitative support for the accuracy of our method, we also directly assessed its accuracy by comparing our CN calls against the results from digital PCR or MLPA. In this direct comparison, 99.8% of our *SMN1* and 99.7% of our *SMN2* CN calls agreed with the digital PCR-based or MLPA-based results. Importantly, our recall is 100% for SMA and 97.8% for carriers, and our precision is 100% for both SMA and carriers.

In this study, we optimized our CN calling to work for individuals of any ancestry and thus limited *SMN1/2* differentiation to the functionally important splice variant plus seven sites in high concordance with the splice variant across all populations (Figure 3A). By quantifying the concordance between all of the reference differences and the splice variant, we were able to identify variations at these sites that, if not accounted for properly (i.e. removed from our analysis) could lead to errors in our CN calls. This would be especially problematic in analyzing Africans because they harbor more diverse haplotypes. Future population genetic studies, possibly including using long read sequencing, will help profile the haplotypic diversity across populations more directly and identify new variant sites that could further improve the accuracy of *SMN1*/*SMN2* differentiation.

An important area for improvement is the detection of “silent” carriers. One type of “silent” carrier occurs when an individual has two copies of the *SMN1* gene but they are both on the same haplotype. A SNP (c.*3+80T>G) has been used to identify individuals that are at an increased risk of being carriers when *SMN1* CN is two but the risk associated with this SNP can vary greatly between studies and populations (Table S7, Table S8). When an individual has just one copy of *SMN1* they can be definitively identified as a carrier, but this variant only indicates a 2-8% chance of being a carrier when *SMN1* CN is two (Table S8). With GS, it would be possible to catalog the different variants that occur with different CN combinations of *SMN1* and *SMN2* and possibly identify additional markers that could be used to improve our ability to identify these “silent” carriers. In addition, the loss of the c.840C splice variant currently explains around 95% of SMA cases and the remaining cases include other pathogenic variants. These other pathogenic variants represent another type of “silent” carrier and as more of them are identified, we will extend this software to directly genotype these as part of the testing process, further improving the ability to detect SMA carriers and cases.

While there exist difficult regions in the genome where normal GS pipelines do not deliver variant calls, here we demonstrate the ability to apply GS paired with a targeted informatics approach to solve one such difficult region. So far, this targeted strategy (GS + specialized informatics) has been applied successfully to a number of difficult variants, such as repeat expansions^38^ and CYP2D6^39^. Traditionally, it is not cost effective to perform all of the known genetic tests and carrier screening on every individual, so candidates for specific genetic testing are identified using information such as the carrier rate and family history. However, this process means that many people without a family history who would benefit from knowing their SMA status do not routinely have access to this data. Once GS analysis can detect all SNVs and CNVs in all clinically relevant genes accurately then a more general and population-wide genetic testing strategy will be feasible with a single test. Improving GS to become economical as a substitute for one current genetic test will help facilitate the integration of more genetic tests and carrier screens into GS, allowing more general access to genetic testing population-wide. GS provides a valuable opportunity to assess the entire genome for genetic variation and the continued development of more targeted informatics solutions for difficult regions with GS data will help bring the promise of personalized medicine one step closer to a reality.

### Software and data availability

The *SMN* copy number caller described here can be downloaded from: https://github.com/Illumina/SMNCopyNumberCaller.

The 1kGP data can be downloaded from https://www.ncbi.nlm.nih.gov/bioproject/PRJEB31736/. Data from the NIHR BioResource participants have been deposited in European Genome-phenome Archive (EGA) at the EMBL European Bioinformatics Institute (accession codes available at Ref 30^30^). Those participants from the NIHR BioResource who enrolled for the 100,000 Genomes Project–Rare Diseases Pilot can be accessed by seeking access via Genomics England Limited following the procedure outlined at: https://www.genomicsengland.co.uk/about-gecip/joining-research-community. The Bam files from the NGC individuals have been deposited in EGA under accession number EGAD00001004357.

### Ethics approval and consent

The 13,343 individuals from the NIHR BioResource Rare Diseases project were recruited through NHS Cambridge University Hospitals Foundation Trust under Cambridge South Research Ethics Committee approval 13/EE/0325.

The 65 patient-derived DNA samples from the Motor Neuron Diseases Research Laboratory were isolated from established fibroblast or lymphoblastoid cell lines. For those cell lines obtained from non-commercial sources, biospecimens were obtained after written consent or assent and parental permission. This study was approved by the Nemours/Alfred I. duPont Hospital for Children Institutional Review Board. These samples were de-identified so that no protected health information is known for these lines.

## Data Availability

The 1kGP data can be downloaded from https://www.ncbi.nlm.nih.gov/bioproject/PRJEB31736/. Data from the NIHR BioResource participants have been deposited in European Genome-phenome Archive (EGA) at the EMBL European Bioinformatics Institute. Those participants from the NIHR BioResource who enrolled for the 100,000 Genomes Project-Rare Diseases Pilot can be accessed by seeking access via Genomics England Limited following the procedure outlined at: https://www.genomicsengland.co.uk/about-gecip/joining-research-community. The Bam files from the NGC individuals have been deposited in EGA under accession number EGAD00001004357.

## Acknowledgements

This work was supported by the Cambridge Biomedical Research Centre and the National Institute for Health Research (NIHR) for the NIHR BioResource (grant number RG65966), the National Institute of General Medical Sciences of the National Institutes of Health (P30GM114736 and P20GM103446; to MERB) and the Nemours Foundation (to MERB). We thank the New York Genome Center (supported by NHGRI Grant 3UM1HG008901-03S1), and the Coriell Institute for Medical Research for generating and releasing the 1kGP GS data. We thank NIHR BioResource volunteers for their participation, and gratefully acknowledge NIHR BioResource centres, NHS Trusts and staff for their contribution. We thank the National Institute for Health Research and NHS Blood and Transplant. The views expressed are those of the authors and not necessarily those of the NHS, the NIHR or the Department of Health and Social Care.

